# National Smoking Rates Correlate Inversely with COVID-19 Mortality

**DOI:** 10.1101/2020.06.12.20129825

**Authors:** Michael J. Norden, David H. Avery, Justin G. Norden, David R. Haynor

**Affiliations:** U of Washington (retired); University of Washington, Emeritus; Stanford University School of Medicine, MD Candidate; University of Washington

## Abstract

**Introduction:** Recent studies show cigarette smokers are markedly under-represented among patients hospitalized for COVID-19 in over a dozen countries. It is unclear if this may be related to confounding factors such as age distribution, access to care, and inaccurate records. We hypothesized that these concerns could be avoided by studying smoking prevalence in relation to COVID-19 mortality. Since climate has been identified as a factor in COVID-19, we studied groups of countries with relatively comparable temperatures.

**Methods:** The 20 hottest and 20 coldest countries in the Johns Hopkins Mortality Analysis database with a minimum mortality rate of .3 deaths/100,000 were selected on the basis of the average temperatures of their largest city. Mortality rates were determined as of May 1, 2020 and correlated with national smoking rate adjusting for sex ratio, obesity, temperature, and elderly population.

**Results:** A highly significant inverse correlation between current daily smoking prevalence and COVID-19 mortality rate was noted for the group of hot countries (R=-.718, p = .0002), cold countries (R=-.567, p=.0046), and the combined group (R=-.324, p=.0207). However, after adjustments only the regression for hot countries and the combined group remained significant. In hot countries, for each percentage point increase in smoking rate mortality decreased by .147 per 100,000 population (95% CI .102-192, p=.0066). This resulted in mortality rates several-fold elevated in the countries with the lowest smoking rates relative to the highest smoking rates. In the combined group, mortality decreased by .257 per 100,000 population (95% CI .175-.339, p=.0034).

**Discussion:** These findings add support to the finding of an inverse relationship between current smoking and seriously symptomatic COVID-19. However, we conclude that the difference in mortality between the highest and lowest smoking countries appears too large to be due primarily to the effects of smoking per se. A potentially beneficial effect of smoking is surprising, but compatible with a number of hypothetical mechanisms which deserve exploration: 1) Studies show smoking alters ACE2 expression which may affect COVID-19 infection or its progression to serious lung pathology. 2) Nicotine has anti-inflammatory activity and also appears to alter ACE2 expression. 3) Nitric oxide in cigarette smoke is known to be effective in treating pulmonary hypertension and has shown in vitro antiviral effects including against SARS-CoV-2. 4) Smoking has complicated effects on the immune system involving both up and down regulation, any of which might alone or in concert antagonize progression of COVID-19. 5) Smokers are exposed to hot vapors which may stimulate immunity in the respiratory tract by various heat-related mechanisms (e.g. heat shock proteins). Studies of steam and sauna treatments have shown efficacy in other viral respiratory conditions. At this time there is no clear evidence that smoking is protective against COVID-19, so the established recommendations to avoid smoking should be emphasized. The interaction of smoking and COVID-19 will only be reliably determined by carefully designed prospective study, and there is reason to believe that there are unknown confounds that may be spuriously suggesting a protective effect of smoking. However, the magnitude of the apparent inverse association of COVID-19 and smoking and its myriad clinical implications suggest the importance of further investigation.

## INTRODUCTION

Smokers are markedly under-represented among hospitalized patients testing positive for COVID-19.^1,2^ This is surprising as smoking is generally associated with greatly exacerbating respiratory infections.^3^ A systematic review of Chinese studies totalling 5960 inpatients showed the pooled prevalence of current smokers was 6.5%, approximately a quarter of the population smoking prevalence.^2^ Researchers in France noting the Chinese data and a U.S. report showing a roughly ten-fold under-representation of smokers among 7171 COVID inpatients and outpatients, were motivated to conduct a cross-sectional study in France.^1^ Among a group of 343 inpatients they found a smoking rate of 4.4%, and among 139 outpatients a rate of 5.3%, each more than four-fold below community rates adjusted for age and sex.

A recent review of smoking and COVID-19 by Simons, et al. found that a similar under-representation was apparent in seven other countries: Italy, Israel, Kuwait, Mexico, Spain, Switzerland, and the U.K.^4^ They found only marginal under-representation in South Korea and a reversed pattern in Iran. Using sex-adjusted rates of smoking prevalence there was moderate under-representation of smokers in South Korea -- 18.5% of 28 hospitalized patients^5^ vs a sex-adjusted national rate of 28.6%,^6^ and in Iran -- 14% of 490 hospitalized COVID-19 patients ^7^ vs. a sex-adjusted national rate of 24.6%.^8^ Additionally, a small study from Germany found a smoking rate of 6% of 50 hospitalized patients^9^ vs. a sex adjusted national rate of 24.9%. ^10^ However, with the exception of the French study and our adjustments for sex-ratio, all of the preceding community rates are not adjusted for demographics. The most critical adjustment is likely age, as COVID-19 inpatients are heavily skewed toward the elderly and smoking prevalence drops precipitously in this age group, falling to an average of 11.5% among those over 65 years old across 17 European countries.^11^

The smoking prevalence among people over age 65 in New York City (NYC) is 9% among men and 7% among women.^12^ These smoking prevalences allow a good comparison to COVID-19 admissions for two New York City hospitals. Current smokers accounted for only 3.5% of the 3273 hospitalized with COVID-19 at Mt. Sinai and only 4.2% of deaths.^13^ Similarly, in the first 1000 patients hospitalized at New York-Presbyterian Co in NYC current smokers accounted for only 4.1% of patients hospitalized for COVID-19 and 4.2% COVID-19 patients treated in the ICU.^14^ Other US data from 2,489 COVID inpatients across 13 states showed 6% current smokers.^15^ Thus, compared to earlier reports, these more recent US data continue to show less striking but still substantial under-representation of smokers among COVID-19 inpatients.

The most extensive study of COVID-19 in the U.K. based on 16,749 inpatients in the initial fully adjusted model found a small under-representation of smokers (OR .88, 95% CI .79-.99). In a post-hoc analysis, however, when further individual factors were added, the OR dropped to .94 and was no longer significant.^16^ The mortality figures were more compelling, of 5683 deaths recorded, only 393 (6.9%) were in current smokers, whereas current smokers constituted 17% of the cohort.

Data sets that include non-hospitalised patients present more mixed results. A prospective study using the UK Biobank identified 908 patients testing positive for COVID, a slight under-representation of smokers in that group relative to those testing negative.^17^ However, after controlling for other variables they found a non-significant over-representation. Similarly, an online survey of 53,002 found smokers slightly over-represented among those reporting positive tests or probable cases OR 1.11 (95% CI .03-1.2), but only for those having less than 16 years of education.^18^ The study also showed smokers to be more worried about contracting COVID-19, but also less compliant with social distancing recommendations. In Mexico of 7497 people testing positive for COVID-19 only 9.4% were current smokers,^19^ vs. a sex adjusted national rate of 18.0%.^20^ Finally, among veterans aged 54-75 years, 585 out of 3789 tested COVID-19 +. Smokers were less likely to test positive (OR .43, 95% CI .35-.57), but there was no significant difference in hospitalization.^21^

The apparent substantial under-representation of smokers among COVID-19 inpatients consistently across thirteen countries is remarkable, but variations in results highlight the importance of considering potential confounds. For the most part, reports are not corrected for age or comorbidity, and moreover the records may be incomplete with regard to smoking status. It is also possible that the patient samples were for some reason non-representative (e.g. including many long-term care patients who are not allowed to smoke, higher socioeconomic classes with better access to healthcare, numerous health-care workers, etc), that patients concealed their smoking habit or were too sick to inform, that they failed to seek help because of misinterpretation of symptoms (“smokers cough”), that changes in their bodies from years of smoking somehow produced false negative COVID-19 test results, or that those smokers most at risk for hospitalization had already died or had quit smoking because of their infirmity (reverse causality). We therefore sought to test the association in a way that was not subject to any of these confounds. Our hypothesis was that if smoking actually was associated with a several-fold reduction of the risk of being hospitalized for COVID-19, there should be some reduction in COVID-19 mortality rates in communities with more smokers -- assuming other confounds are controlled.

However, many countries of Western Europe had not only high smoking prevalence but some of the highest COVID-19 mortality, so any comparisons with these countries would be unlikely to show such a relationship. Analyses from MIT and the University of Maryland showed most cases of COVID-19 developed within a narrow temperature range.^22,23^ Consistent with this we observed that a group of 19 countries stood out in terms of their mortality rates -- all had rates 50% or more above the others. All but two of these countries fell within a relatively narrow band of temperatures (over the January to April period) - bounded by the temperatures of Austria on the low end (6.75 degrees C) and Portugal on the high end (13 degrees C). We hypothesized that if there was a protective effect of smoking it might be possible to detect it outside of this moderate temperature band where temperature appeared to be a dominant factor and mortality rates were extreme.

## METHODS

To investigate the relationship between national smoking prevalence and COVID-19 mortality we chose two cohorts: “hot countries” and “cold countries.” All countries available in the Johns Hopkins mortality analysis^24^ as of 5/1/20 with mortality rates of at least .3 deaths/100k population were included in the analysis. A minimum mortality threshold was required because extremely low mortality rates may reflect inadequate testing — furthermore, this limits the impact of floor effects in the analysis. Mortality rates were chosen as the main outcome measure, as this reflects not only COVID-19 deaths but the preceding hospitalizations, both of which could potentially be decreased in smokers. All 20 countries colder than Austria were included in the cold countries cohort. An equal size group was formed for the 20 hottest countries.

## Data Sources

### Mortality data

National mortality figures, Deaths/100,000 population, were obtained for each country in the study from the Johns Hopkins Mortality Analysis on May 1, 2020.^24^

### Temperature Data

The months January through April were chosen for temperature comparisons as they are the months preceding our mortality statistics and include the period when the virus was known to be spreading outside of China. Monthly temperature averages for the largest city in each country/state were obtained from the website timeanddate.com. The largest city was selected as the reference to establish a rough approximation of the temperature experienced by the whole of the country/state population.

### Sex Ratio

The number of males per 100 females for each country was obtained from statisticstimes.com — Original source: UN (World Population Prospects 2019)/World Bank.^25^

### Obesity Prevalence

National obesity rates, defined as a Body Mass Index (BMI) over 30, were obtained from worldpopulationreviews.com.

### Percentage of elderly in the population

National data was obtained for percentage of the population over 65 years of age from worldbank.org.

### Percentage of daily smokers

Male and female percentages were obtained for each country from tobaccoatlas.org^26^ and sex ratio was used to calculate overall percentages. Note that daily smoking prevalence will be lower than overall smoking prevalence which is more often cited. Daily smoking was chosen because there might be a dose effect and daily smokers would be the most affected.

## Analyses

Statistical analyses were performed using the R programming language version 3.5.3. For each of the groups, and for the combination, a linear correlation analysis was performed evaluating the relationship between daily smoking rates and COVID-19 mortality. Second, for each of the groups multiple regression was performed to adjust for known risk factors for COVID-19 mortality. The independent variables in the regression were daily smoking prevalence, population percentage over 65, sex ratio, obesity prevalance, and average ambient temperature from January-April, 2020. Post hoc analyses were run with and without inclusion of Korea and Sweden, because of their unique COVID-19 policies and procedures.

## RESULTS

Correlations and scatterplots between national smoking prevalence and COVID-19 mortality rates in each of the three groups of countries can be seen below in Figure 1 (Hot Countries), Figure 2 Cold Countries) and Figure 3 (Hot and Cold Countries Combined).

**Figure 1.**
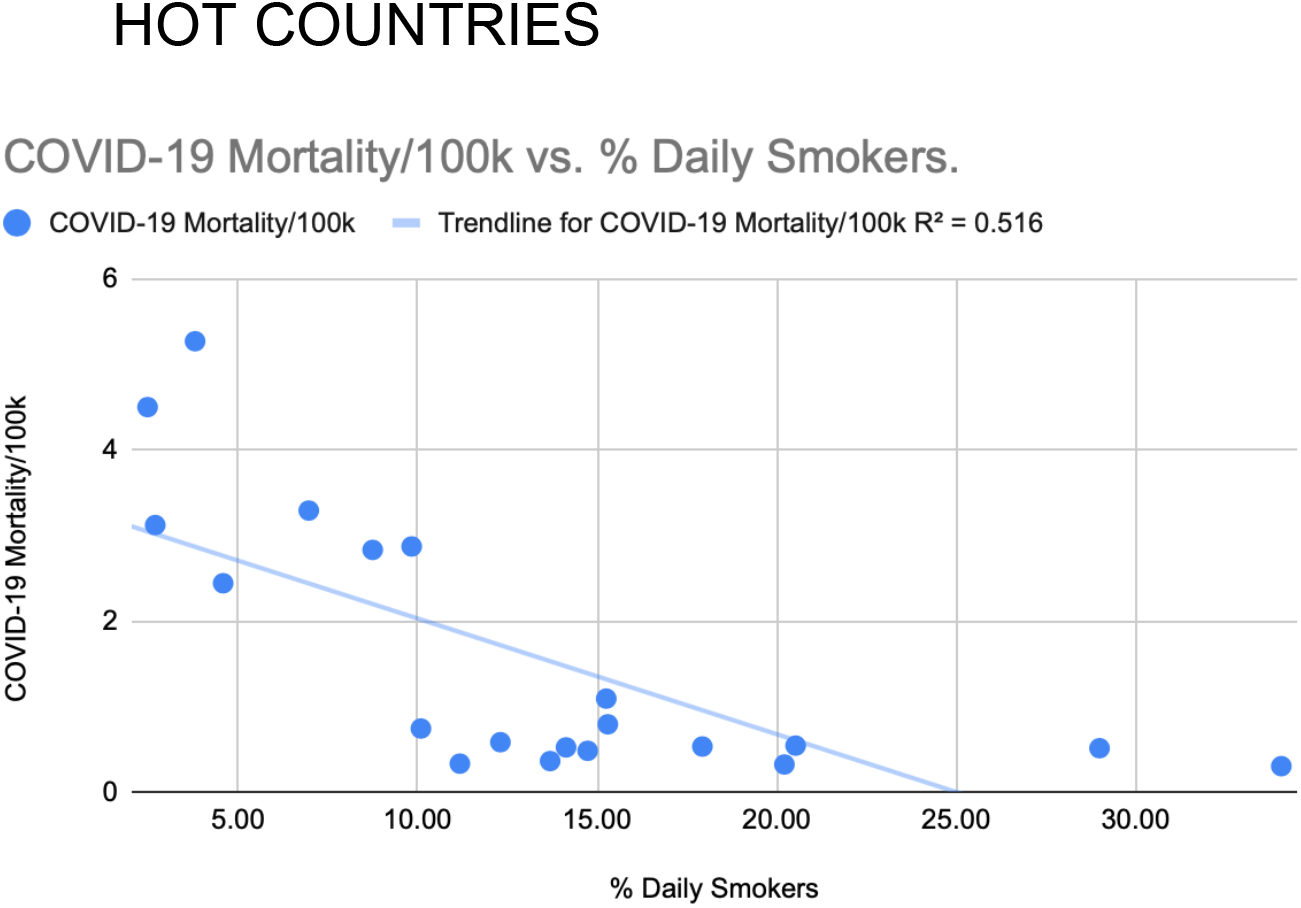
Daily smoking prevalence correlated inversely with national COVID-19 mortality rates of the 20 hottest countries. Pearson’s correlation without adjustments: R=-.718, p=.0002.

**Figure 2.**
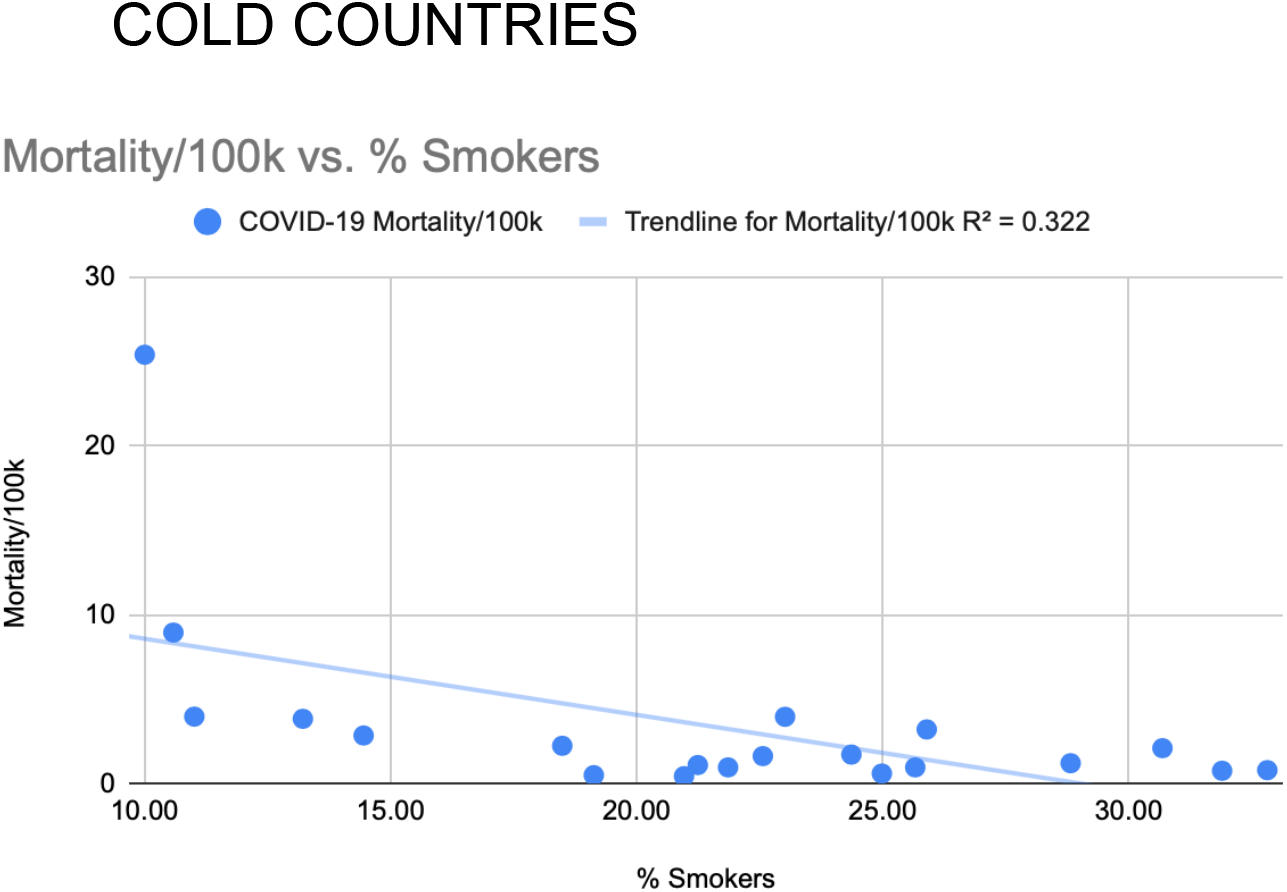
Daily smoking prevalence correlated inversely with national COVID-19 mortality rates of the 20 coldest countries. Pearson’s correlation without adjustments: R=-.567 p=.0046

**Figure 3.**
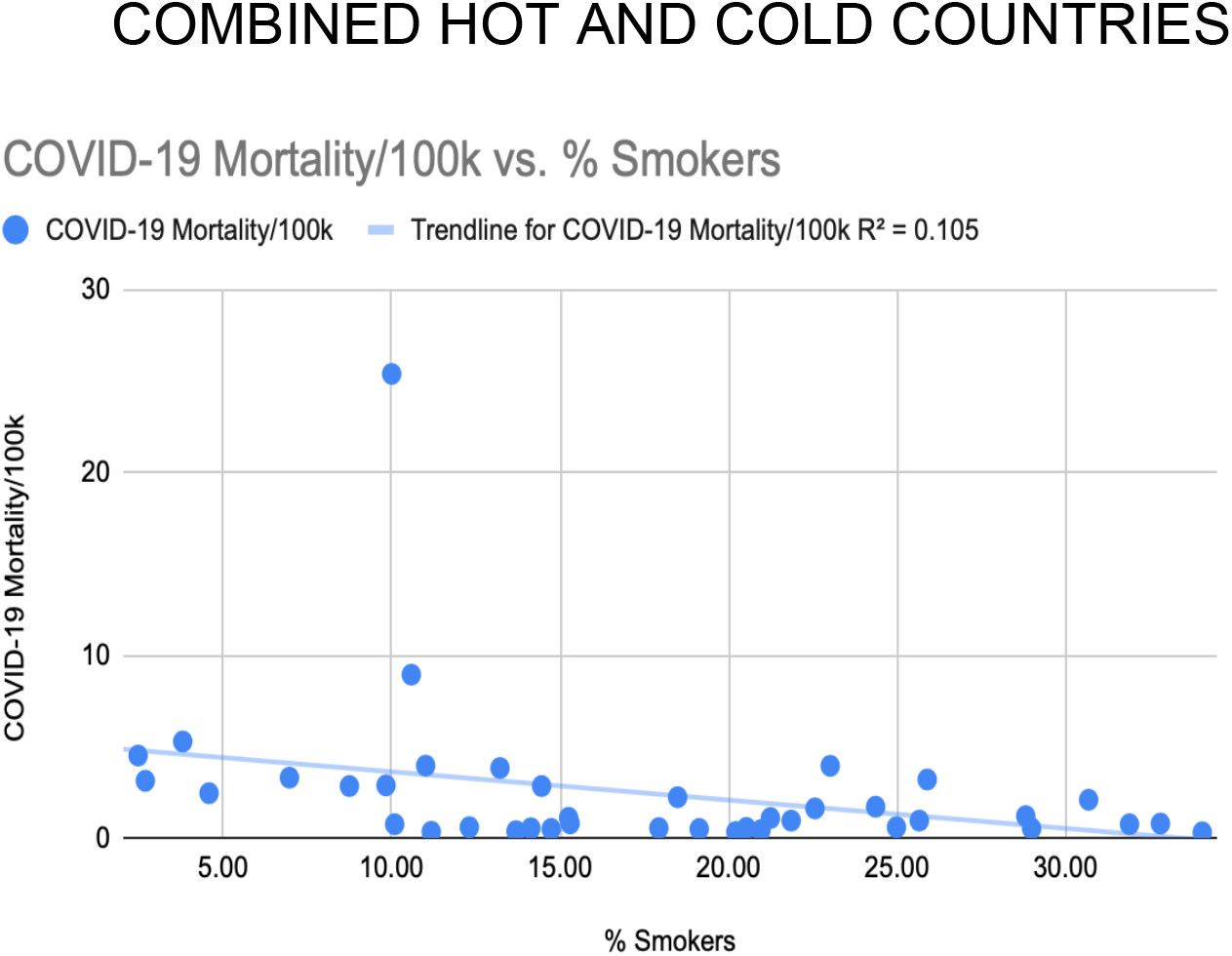
Daily smoking prevalence correlated inversely with national COVID-19 mortality rates for the combined group of the 20 hottest countries and the 20 coldest countries. Pearson’s correlation without adjustments: R=-.324, p=.0207.

Table 1 shows the estimates of the linear regression model coefficients evaluating the relationship between daily smoking prevalence and COVID-19 mortality rates incorporating the studied variables. For hot countries each percentage point increase in smoking prevalence caused a COVID-19 mortality decrease of .147 per 100,000 (95% CI .102-.192, p=.0066). For cold countries there was no significant relationship seen between smoking prevalence and COVID-19 mortality when including the confounding variables (model coefficient = .066, 95% CI -.174-.042, p=.5526). For the combined group of 40 for each percentage point increase in smoking prevalence, COVID-19 mortality decreased by .257 per 100,000 (95% CI .175-.339 p=.0034).

**Table 1.** Regression model coefficients on mortality per 100,000 people.

**Hot Countries**
|  | Estimate | Std. Error | t value | Pr(>\|t\|) |
| --- | --- | --- | --- | --- |
| (Intercept) | -0.6164 | 5.0715 | -0.1215 | 0.9053 |
| Smoking % | -0.1468 | 0.0448 | -3.2755 | **0.0066** |
| Temperature | 0.1459 | 0.1587 | 0.9193 | 0.3760 |
| Obesity % | 0.0183 | 0.0568 | 0.3230 | 0.7522 |
| Age > 65 y.o. | 0.0180 | 0.0968 | 0.1862 | 0.8554 |
| Sex Ratio | -0.0014 | 0.0151 | -0.0910 | 0.9290 |

**Cold Countries**
|  | Estimate | Std. Error | t value | Pr(>\|t\|) |
| --- | --- | --- | --- | --- |
| (Intercept) | -14.1803 | 13.2526 | -1.0700 | 0.3075 |
| Smoking % | -0.0661 | 0.1078 | -0.6126 | **0.5526** |
| Temperature | -0.3094 | 0.3341 | -0.9261 | 0.3742 |
| Obesity % | 0.1619 | 0.0949 | 1.7053 | 0.1162 |
| Age >65 y.o | 0.0575 | 0.1548 | 0.3716 | 0.7173 |
| Sex Ratio | 0.1554 | 0.1191 | 1.3049 | 0.2186 |

**All Countries**
|  | Estimate | Std. Error | t value | Pr(>\|t\|) |
| --- | --- | --- | --- | --- |
| (Intercept) | 4.6025 | 5.6480 | 0.8149 | 0.4210 |
| Smoking % | -0.2572 | 0.0816 | -3.1525 | **0.0034** |
| Temperature | -0.0855 | 0.1035 | -0.8261 | 0.4147 |
| Obesity % | -0.0530 | 0.1091 | -0.4860 | 0.6302 |
| Age >65 y.o | 0.2434 | 0.2016 | 1.2074 | 0.2359 |
| Sex Ratio | 0.0166 | 0.0225 | 0.7360 | 0.4669 |

To test whether any outliers may be driving the results we performed post-hoc analyses removing Sweden and Korea both individually and together. Sweden’s policy of minimal mandatory closures of schools and business appeared to result in greatly elevated mortality rates. In contrast, South Korea’s aggressive testing, contact-tracing, and utilization of masks appeared to result in an exceptionally low mortality rate. Removing either or both increased the inverse correlation in each subgroup and the combined groups. Removing both countries from the combined group did not change the result - each percentage point increase in daily smoking prevalence, COVID-19 mortality decreased by .15 per 100,000 (95% CI .115-.181, p=9.22E-05).

## DISCUSSION

A significant inverse correlation of smoking prevalence and COVID-19 mortality rate was found for the subgroups of hot and cold countries, and for the combined subgroups. After adjustments were made for age (% of the population > 65 y.o.), sex ratio, temperature (January - April average), and obesity percentage, the effect of smoking prevalence remained significant for the hot countries, and the combined subgroups, but not for the cold countries. The association was stronger in the post hoc analysis for the combined groups excluding the outliers Korea and Sweden. It must always be kept in mind that correlation does not imply causation, and although we found the surprising hypothesized correlations supporting a protective role of smoking in COVID-19, we now question if this association might instead reflect an unknown confound.

The correlation found, especially in the hot countries, appears too strong to be based primarily upon an effect of smoking. Post hoc, it is apparent by inspection of the scatterplots that countries with the lowest smoking prevalence have several-fold higher mortality rates relative to the lowest smoking countries. This cannot be the result of smoking per se. The reason is that even if we assume every smoker is 100% protected from developing COVID-19, there are too few smokers in the population to produce such a large effect, and it is reasonable to assume that there is a confounding influence. For this reason, although the current study reinforces a strong inverse association, at the national level, of smoking and seriously symptomatic COVID-19, these data suggest that this association is, at least in large part, based on something other than smoking itself.

This same interpretation may apply to a study by Gonzalez-Marron and Martinez-Sanchez,^27^ which appeared in preprint form while we were preparing this manuscript. Similarly, they found a significant inverse association of smoking with national per capita rates of COVID-19 cases in the 27 countries of the European Union (EU). They found no significant association with the national case fatality rate, but they did not look at the national mortality rate. Since most of the countries in the EU have a temperate climate, this suggests that an inverse association of smoking and COVID-19 is not limited to extreme climates. However, when we calculated from their data the case rates in their seven highest smoking and seven lowest smoking EU countries (approximately quartiles), we similarly saw a nearly three-fold difference in median rates (41.8 vs. 112.14). Again, such a large difference does not appear compatible an effect of smoking itself, and implicates a powerful confounding influence.

What this confound or confounds might be is unclear. Countries with high smoking prevalence may generally have weaker health care systems with less access to treatment and, importantly, less testing. More generally, smoking prevalence may be linked to myriad national differences in political structures, economics, or behavioral tendencies (e.g. international travel) that impact the acquisition, diagnosis, treatment, or reporting of COVID-19. Additionally, patients or physicians may assume that the smokers’ pulmonary symptoms are a result of smoking and not test for COVID-19. This may be especially true when testing capabilities are limited. Finally, in our study one confound could be that COVID-19 deaths occurring in smokers may frequently not be counted because of smokers dying in the community undiagnosed with COVID-19 -- something which would happen more often in a country with a poor healthcare system and limited testing.

The inverse correlation may, of course, have both physiologic and nonphysiologic underpinnings. The strongest case for physiology appears to be the substantial under-representation of smokers among hospitalized patients that has been consistently observed across thirteen countries. If smoking acts in some way to reduce COVID-19 mortality, it seems that this must be occurring prior to hospitalization, as data show hospitalized smokers are much more likely to experience serious progression of illness relative to non-smokers.^28,29^ However, it is hard to reconcile this consistent pattern with the results of the Simon’s review showing little evidence of under-representation of smokers among those testing COVID-19 positive in the community, or evidence that smokers testing positive are less likely to be hospitalized -- though here results were mixed.^4^ So the under-representation of smokers among those hospitalized remains puzzling.

To our knowledge, the only other respiratory virus for which smoking has been suggested to have a protective effect is the closely related SARS-CoV. A case-study was conducted in 2003 to investigate this possibility. While it was found that smokers were more than four-fold under-represented in this group of SARS patients, when confounds were considered this dropped to only 1.7-fold -- and the effect was not significant.^30^ This under-powered study concluded that “smoking is shown to provide no protection,” yet intriguingly there was clearly some suggestion of association in a closely related virus.

If smoking actually confers some protection from seriously symptomatic COVID-19, what are some possible mechanisms of action? There is evidence that smoking affects the ACE-2 receptors used by the virus for cell entry, but studies are mixed whether it upregulates or down regulates this receptor.^31-33^ While down regulation can easily be understood to potentially be protective from infection, up-regulation might be protective from development of severe lung disease. In the case of SARS-CoV, a study showed it down-regulates the ACE2 and that this was directly implicated in the development of severe lung disease.^34^

A group in France proposed a potential role of nicotine as the agent responsible for smoking’s action on the ACE2 receptor, potentially providing a protective mechanism for smoking in COVID-19. Nicotine is also of interest because it could help in COVID-19 by mitigating a host of destructive inflammatory reactions^35,36^ that often are associated with deterioration in the later stages of COVID-19.^37,38^

Another mechanism of interest is nitric oxide (NO) which is one of the many chemicals found in cigarette smoke, and also relevant to nicotine. Smokers are exposed to high levels of NO in the inhaled smoke as well as endogenously released NO after uptake of nicotine in the brain.^39^ Nitric oxide has antiviral activity, and has been shown to reduce infectivity of H1N1 in vitro.^40,41^ In colds, some of which are caused by coronaviruses,^42^ increased nitric oxide is associated with more rapid viral clearance.^43^ Evidence also points to antiviral activity with SARS-CoV. There has been an anecdotal report of successful administration of gaseous NO in COVID-19.^44^ Two multicenter randomized controlled trials are planned, organized by investigators at Massachusetts General Hospital in Boston. These trials are aimed primarily at treating acute respiratory distress in COVID-19 patients.^45^

Smoking is associated with inflammation in the respiratory tract and creates a host of changes to the immune system, and it is possible that some of these changes might make for an inhospitable environment for SARS-CoV-2 infection. A comprehensive review of immune changes associated with smoking concluded that these effects were almost always harmful rather than beneficial.^46^ As noted, the exceptions may involve SARS-CoV and SARS-CoV-2, and it could be informative to understand what is so different about how these viruses act, and to develop treatments based on this understanding.

Lastly, heat could be the basis of a protective mechanism directly to the warm vapors being inhaled. The internal temperature of a cigarette averages around 688 degrees C.^47^ Smoke in the mouth can reach temperatures of 60 degrees.^48^ Heat may be therapeutic in a number of ways. Fever is an evolutionarily conserved function in all vertebrates,^49^ it’s generally believed to play a role in the body’s defense against viruses,^50,51^ and enhances a host of immune functions, including productions of heat shock proteins,^50,51^ Importantly, COVID-19 patients that are admitted to the hospital without fever have been found to have greatly elevated risk of poor outcomes.^29,52,53^ Furthermore, we know that coronaviruses are highly temperature sensitive — at least in the lab where virus inactivation was reduced from taking 14 days at 4 degrees C to five minutes at 70 degrees C.^54^ Aside from the warm smoke vapors inhaled, there is also another way that heat relates to smoking. A recent study showed that smokers have a full one degree C elevated breath temperature compared to non-smokers.^55^ Furthermore, in the hour after a cigarette is smoked breath temperature is further increased by an average of .42 degrees C. Smokers are chronically experiencing a localized relative heating of the respiratory tract, believed to be principally related to increased blood flow associated with inflammation.^56^

To the extent that temperature is implicated in a smoker’s physiology, it is reasonable to consider that there are a host of much more benign ways to warm the respiratory tract than smoking. Though the evidence was deemed inconclusive in a Cochrane review,^57^ several controlled trials^58-60^ were cited of various heat treatments (e.g. steam) showing successful reduction of symptoms or duration of colds, another illness sometimes caused by a coronavirus.^61^ Another obvious treatment to investigate is the sauna, as there are similarly promising preliminary evidence of efficacy in colds.^62^ Furthermore, sauna use has the additional benefit of regular usage being associated with huge reductions of some of the major comorbid risk factors associated with COVID-19, a 40% reduction of respiratory illness (including pneumonia)^63^ and a 50% reduction in cardiovascular illness.^64^ Finally, a study is planned at UT Southwestern Medical Center to treat COVID-19 patients undergoing mechanical ventilation with esophageal heat exchangers.^65^ COVID-19 patients at the point of needing ventilation are typically without fever, and the goal of this treatment is to bring their body temperature up to a maximum of about 39 degrees C.

The primary strength of the current study is that it shows a strong inverse association between COVID-19 mortality and daily smoking prevalence not subject to the many confounds identified in previous studies reporting under-representation of smokers among COVID-19 patients. It is, of course, still subject to other potential confounds, and the inverse association between smoking and COVID-19 mortality we found did not hold up to full adjustments for one of the two cohorts analyzed individually. The current study has other limitations, while it uses COVID-19 mortality rates that are likely more accurate than case rates, they nonetheless still depend in on testing availability and accuracy. Additionally, all reported national statistics are also subject to political influences, lockdown variability, regional infection rates, timing of initial cases Furthermore, this is still an evolving pandemic and mortality rates will change. At this time there is no clear evidence that smoking is protective against COVID-19, so the established warnings to avoid smoking should be emphasized. The interaction of smoking and COVID-19 only be reliably determined by carefully designed prospective study, and there is reason to eve that there are unknown confounds that may be spuriously suggesting a protective effect smoking. However, the magnitude of the apparent inverse association of COVID-19 and smoking and its myriad clinical implications suggest the importance of further investigation.

## Data Availability

All data in the paper is publicly available and the corresponding author would be happy to assist interested researchers. Listed below are the links, which are also included in the paper, and listed in the next section for external data:
https://coronavirus.jhu.edu/map.html
https://timeanddate.com
https://statisticstines.com
https://worldpopulationreviews.com
https://worldbank.org
https://tobaccoatlas.org

https://coronavirus.jhu.edu/map.html

https://timeanddate.com

https://statisticstines.com

https://worldpopulationreviews.com

https://worldbank.org

https://tobaccoatlas.org

